# Cross reactivity of spike glycoprotein induced antibody against Delta and Omicron variants before and after third SARS-CoV-2 vaccine dose

**DOI:** 10.1101/2021.12.30.21268308

**Authors:** Sian E Faustini, Adrian M Shields, Gemma Banham, Nadezhda Wall, Saly Al-Taei, Chloe Tanner, Zahra Ahmed, Elena Efstathiou, Neal Townsend, Tim Plant, Marisol Perez-Toledo, Aleksandra Jasiulewicz, Ruth Price, James McLaughlin, John Farnan, Julie Moore, Louise Robertson, Andrew Nesbit, Grace Curry, Amy Black, Adam F Cunningham, Lorraine Harper, Tara Moore, Mark T Drayson, Alex G Richter, On behalf of the COVID-HD Birmingham study group and the PITCH consortium

## Abstract

Variants of SARS-CoV-2 may evade natural and vaccine induced immunity and monoclonal antibody immunotherapeutics. There is an urgent need to know how well antibodies, induced by healthy and Clinically Extremely Vulnerable (CEV) patients, will bind and thus help reduce transmission and severity of infection from variants of concern (VOC). This study determines the cross-reactive binding of serum antibodies obtained prior to and 28 days after a third vaccination in three cohorts; a health care worker cohort who received three doses of Pfizer-BioNtech (PPP), a cohort of CEV patients received two doses of the AstraZeneca-ChAdOx1-nCoV-19 (AAP) vaccine, followed by a third PFZ vaccine and a haemodialysis cohort that had a mixture of two AZ or PFZ vaccines followed by a PFZ booster. Six months post second vaccine there was evidence of antibody waning with 58.9% of individuals in the HD cohort seropositive against Wuhan, 34.4% Delta and 62.2% Omicron strains. For the AAP cohort, equivalent figures were 62.5%, 45.8% and 91.7% and the PPP cohort 92.2%, 90% and 91.1%. Post third dose vaccination there were universal increases in seropositivity and median optical density. For the HD cohort, 98.8% were seropositive to the Wuhan strain, 97.6% against Delta and 100% against Omicron strains. For the PPP and AAP cohorts, 100% were seropositive against all 3 strains. Lastly, we examined the WHO NIBSC 20/136 standard and there was no loss of antibody binding to either VOC. Similarly, a dilution series of Sotrovimab (GSK) found this therapeutic monoclonal antibody bound similarly to all VOC.

**Highlights:**

- IgG anti-SARS-CoV-2 Omicron spike glycoprotein antibody levels were high in 100% of health care workers (HCW), a general practice population considered clinically extremely vulnerable (CEV) and haemodialysis patients (HD) 4 weeks after a third SARS-CoV-2 vaccine dose (Pfizer-BioNtech-PFZ).
- For both Delta and Omicron variant spike glycoproteins these antibody levels were highest in the CEV cohort who had previously received two doses of AstraZeneca ChAdOx1 nCoV-19 vaccine (AAP), lower in HCW who had previously received two doses of PFZ (PPP) and lowest in HD who had a mix of vaccines for the first and second dose
- Prior to this third vaccine dose and 6 months post second vaccine dose there was evidence of significant waning of antibodies against VOC.

## Introduction

The Severe Acute Respiratory Syndrome Coronavirus-2 (SARS-CoV-2) pandemic continues to affect almost every country in the world. Variants of SARS-CoV-2 may evade natural and vaccine-induced immunity and thus continuously reset our progress in mitigating the disastrous impacts of the virus on our health and socioeconomic well-being. In the UK, two main vaccines have formed the basis of the national immunisation strategy: the AstraZeneca ChAdOx1 nCoV-19 vaccine (AZ) (1) and the Pfizer-BioNtech COVID-19 vaccine (PFZ)(2). Both elicit immune responses targeting the original, wildtype SARS-CoV-2 (Wuhan) spike glycoprotein and both have been shown to reduce the incidence of severe disease in clinical trials (1,2). With the emergence of novel variants of concern (VOC) that demonstrate increased transmissibility (3,4), it is necessary to understand whether current vaccines will provide protection against further novel VOC. In mid-2021 the Delta VOC became the dominant strain worldwide and currently the Omicron VOC is likely to becoming the dominant SARS-CoV2 strain in the UK and across the world (5). Indeed, both strains may even continue to co-exist within the same population. These rapid shifts in the pre-dominance of VOC outpaces and impairs the development and testing in clinical trials of new VOC-tailored vaccines. We do not yet know how well the different vaccine strategies applied in the UK will reduce the transmission of and severity of disease arising from rapidly emerging VOC in the general population and immunological vulnerable subgroups.

After vaccination, detection of serum IgG against the SARS-CoV-2 spike glycoprotein correlates with future protection against COVID-19 infection (6). Nevertheless, it is not fully understood whether IgG binds as similarly to the spike glycoprotein from currently circulating VOC as to the vaccine encoded spike glycoprotein and therefore likely to offer protection against infection. This knowledge gap is especially pronounced for immunocompromised or other clinically extremely vulnerable (CEV) patients such as haemodialysis (HD) patients. In this study, we have designed and utilised three ELISA tests to measure IgG levels against SARS-CoV-2 spike glycoprotein; identical except for the use of spike glycoprotein from the original Wuhan strain or from the B.1.617.2 (Delta) or B.1.1.529 (Omicron) VOC. We have used these ELISAs to determine cross reactivity of spike glycoprotein induced antibody against Delta and Omicron variants before and after third SARS-CoV-2 vaccine dose. These results show the value of the additional vaccination on enhancing IgG responses to the spike glycoprotein from both VOCs, even in HD and CEV patients.

## Methods

### Cohorts

We have used serum from three well characterised cohorts. Firstly a health care worker cohort from University Hospitals Birmingham from the *Determining the immune response to SARS-CoV-2 infection in convalescent health care workers (COCO)* study (7,8), who received three doses of PFZ. Secondly, individuals attending general practice for vaccination in Ulster who were considered Clinically Extremely Vulnerable (CEV) as part of The Pandemic Study, who received two doses of the AZ vaccine, followed by a third immunization of the PFZ vaccine. Lastly, individuals on haemodialysis under renal care at the University Hospitals Birmingham that had a mixture of two AZ vaccines or two PFZ vaccines followed by a PFZ booster (9). Samples were taken at 6 months following their 2^nd^ vaccination of their primary course, prior to the 3^rd^ booster vaccination with PFZ and also 28 days following vaccination.

### ELISAs

Here we have used the core design of an anti-IgG/A/M SARS-CoV-2 ELISA (10,11) to measure IgG antibodies specific for spike protein from the original Wuhan strain (Abingdon Health, York Biotech Campus Sand Hutton, York) (12), B.1.617.2 (Delta -Abingdon Health) and B.1.1.529 (Omicron - SinoBiological China). Plates were coated with spike glycoproteins at a concentration of 1 ug/mL onto high binding ELISA plates overnight at 4°C. Plates were washed with PBS-Tween20 and then blocked with 2% BSA diluted in PBS Tween (0.1% Tween-20) for 1 hour at room temperature (RT). Serum samples were diluted at 1:40 with 2% BSA + 0.1% PBS-Tween and added to the plate (100 ul) after washing and incubated for 1 hour at RT. HRP-conjugated mouse-anti-human R10-IgG (monoclonal antibody obtained from Dr. Margaret Goodall, Clinical Immunology Service, University of Birmingham) (1:8000 dilution) was then added to the plate and incubated for 1 hour at RT. Plates were washed and then TMB substrate was used for development. 0.2M H_2_SO_4_ was added to the well after 10 minutes in order to stop the reaction. Optical densities were measured at 450 nm on the Dynex Dynaread (Aspect Scientific). To establish a threshold for seropositivity a set of historic samples were run from pre-2019 which were pre-pandemic and therefore true negatives (n=73).

In addition we ran serial dilutions at 1 in 2 (250 – 7.8 IU/mL) of the WHO standard NIBSC 20/136 (13). We also ran the therapeutic monoclonal antibody therapy Sotrovimab (Glaxo Smith Kline) in serial dilutions. A therapeutic dose of 500mg is given to patients at a concentration of which is 62.5 mg/ml.

### Ethical approval

The COCO study was ethically approved for this work by the London - Camden and Kings Cross Research Ethics Committee on behalf of the United Kingdom Health Research Authority – reference 20/HRA/1817. The Haemodialysis study was ethically approved for this work by the North West-Preston Research Committee on behalf of the United Kingdom Health Research Authority for the National Institute of Health Research Coronavirus Immunological Analysis study – reference 20/NW/ 0240. The Ulster study was ethically approved for this work by the Office of Research Ethics Committee for Northern Ireland on behalf of the United Kingdom Health Research Authority - reference 20/WM/0184. The pre-2019 health controls was ethically approved for this work by the University of Birmingham Research Ethics Committee, United Kingdom - reference ERN_16-178 (2002/201, Amendment Number 4).

### Statistics

All analyses were undertaken on GraphPad Prism 9 (San Diego, California). Normality testing was performed by Kolmogorov –Smirnov testing and data was non parametric. Groups were compared using a Kruskal-Wallis test with Dunn’s multiple comparison test. Statistical significance was accepted when p is less than 0.05.

## Results

### Study demographics

The HD cohort had an average age of 66 years (range 55-75) with male gender predominance (61%). Of this cohort 70.3% had AZ as their primary course and the remaining 29.7% PFZ. There were 90 pre 3^rd^ dose samples tested and 85 samples from those receiving a third vaccination. For the COCO health care worker cohort that had 3 PFZ vaccine courses (PPP) the median age was 47 (range 25-64). Male gender accounted for 21% of this cohort. Ninety pre 3^rd^ dose samples were tested and 60 from individuals vaccinated a third time. The CEV cohort from general practice had a median age of 51 years (range 28-89). Forty four percent of the cohort were male gender. This cohort had two AZ vaccines as their primary course and then a third vaccination with PFZ (termed the AAP cohort). There were 24 pre 3^rd^ dose samples tested and 47 post third vaccination samples tested.

### ELISA results

Testing of sera taken 6 months post second vaccine showed that 58.9% of individuals in the HD cohort were seropositive against the Wuhan strain, 34.4% against Delta and 62.2% against Omicron strains. For the PPP cohort of healthy individuals, 92.2% of individuals remained seropositive against the Wuhan strain, 90% against Delta and 91.1% against Omicron strains. For the AAP cohort of CEV patients, equivalent figures were 62.5% against the Wuhan strain, 45.8% against Delta and 91.7% against Omicron strains (figure 1).

**Figure 1:**
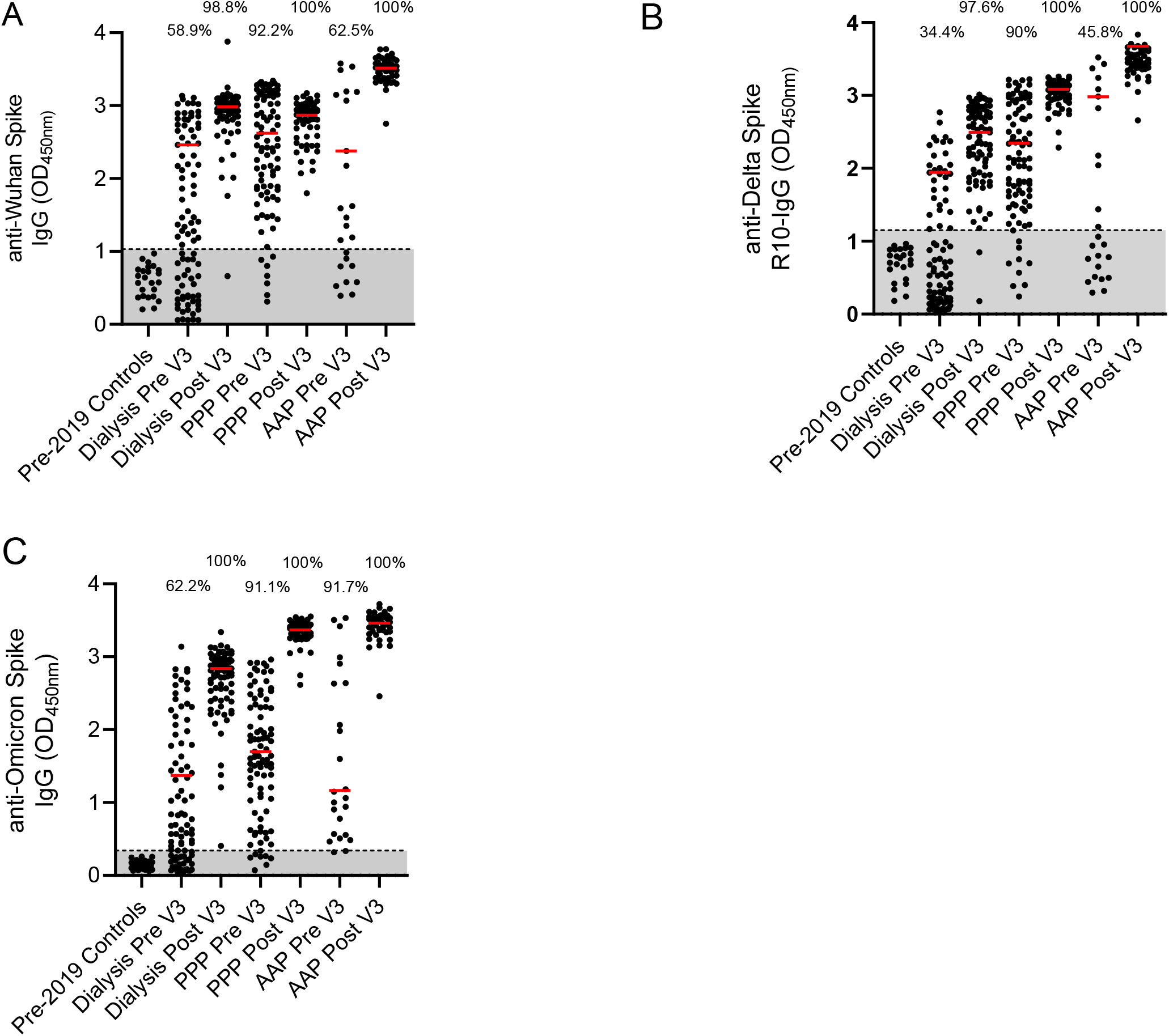
Percentage of cohort with antibodies against Wuhan, Delta and Omicron strains A – Detection of anti-Wuhan spike IgG in pre-2019 controls, a dialysis population, a cohort of health care workers who have had three PFZ vaccines and a Clinically Extremely Vulnerable population in general practice who have had two AZ and one PFZ vaccine. Results are given for pre and post 3^rd^ dose of vaccination. Percentage of cohort that are considered seropositive are included above the dot plots. The red line represents the median of the seropositive individuals in that cohort. B – Detection of anti-Delta for the same populations. C – Detection of anti-Omicron for the same populations Pre – pre 3^rd^ dose of vaccine and 6 months post 2^nd^ dose. Post – 28 days post 3^rd^ dose of vaccine. PPP – 3 Pfizer-BioNtech vaccines given in this cohort. AAP – two AstraZeneca ChAdOx1 nCoV-19 vaccines and the one Pfizer-BioNtech vaccine given in this cohort

Post 3^rd^ dose vaccination a significant increase in seropositivity was observed in the HD cohort and increases in the median optical density (OD) seropositive individuals in all cohorts against spike proteins from viral VOC. For the HD cohort, the proportions of sera that were seropositive were 98.8% to the Wuhan strain, 97.6% against Delta and 100% against Omicron strains. For the PPP and AAP cohorts, 100% were seropositive against all 3 strains (table 1). The median ODs for the HD patients to the Wuhan strain changed from 2.46 to 2.98 (p<0.0001), for Delta 1.94 to 2.49 (non-significant (ns)) and to the Omicron strain 1.37 to 2.84 (p<0.0001). The median OD for the PPP cohort for the Wuhan strain changed from 2.62 to 2.88 (ns), for the Delta strain 2.34 to 3.08 (p<0.0001) and the Omicron strain 1.71 to 3.37 (p<0.0001). For the AAP cohort, the equivalent changes were 2.38 to 3.52 (p<0.0001), 2.99 to 3.45 (p<0.0044) and 1.17 to 3.46 (p<0.0001) respectively.

**Table 1.** SARS-CoV-2 strain.

| SARS-CoV-2 strain | Wuhan |  | Delta |  | Omicron |  |
| --- | --- | --- | --- | --- | --- | --- |
|  | Seropositive (%) | Median OD of seropositive individuals | Seropositive | Median OD of seropositive individuals | Seropositive | Median OD of seropositive individuals |
| Pre-2019 Controls | 0 | n/a | 0 | n/a | 0 | n/a |
| Dialysis Pre V3 | 58.9 | 2.46 | 34.4 | 1.94 | 62.2 | 1.37 |
| Dialysis Post V3 | 98.8 | 2.98 | 97.6 | 2.49 | 100 | 2.84 |
| PPP Pre V3 | 92.2 | 2.62 | 90 | 2.33 | 91.1 | 1.71 |
| PPP Post V3 | 100 | 2.88 | 100 | 3.08 | 100 | 3.37 |
| AAP Pre V3 | 62.5 | 2.38 | 45.8 | 2.98 | 91.7 | 1.17 |
| AAP Post V3 | 100 | 3.52 | 100 | 3.45 | 100 | 3.46 |

We compared the antibody concentration, defined as median optical density (OD), between the PPP and AAP groups and found the median ODs were consistently higher in the AAP cohort for the Wuhan strain (2.88 v 3.52, p<0.0001) and Delta (3.08 v 3.45, p=0.0022) but not Omicron strains (3.37 v 3.46, p>0.9999).

The WHO NIBSC 20/136 standard was run as a standard curve against the Wuhan, Delta and Omicron spike glycoproteins by ELISAs and there was no loss of antibody binding to either the Delta or Omicron VOC. Similarly, a dilution series from 6.5mg/ml to 0.003 mg/ml of Sotrovimab (GSK) found this monoclonal antibody bound similarly to the Omicron spike glycoprotein as to the other spike glycoproteins (Supplementary Figure 1a and b).

## Discussion

Understanding whether antibodies induced to existing vaccines or strains of SARS-CoV-2 can bind and function against novel SARS-CoV-2 VOC and their enhancement or otherwise after additional immunisations, is of critical importance in guiding public health policy during the ongoing SARS-CoV-

2 pandemic. Coupled with this is the necessity to understand how well these responses are sustained over time, whilst de novo vaccines are developed. This knowledge is of particular relevance for clinically vulnerable groups who are typically immunocompromised and either do not make robust antibody responses to vaccination or fail to retain antibody responses over time.

Consistent with early data for healthy adult populations, studies already available show that sera from individuals vaccinated twice binds and neutralises omicron less well than sera from individuals immunized three times (14-16). A distinct feature of the current study is the demonstration that in a cohort of haemodialysis (HD) patients that there is significant waning, 6 months post second dose, of vaccine-induced antibodies against the original vaccine-strain spike glycoprotein (Wuhan) and the Delta and Omicron VOC in comparison to healthy controls. This population of patients are at greater risk of severe SARS-CoV-2 infection, are difficult to shield from infection as they need to attend hospital routinely and also can show diminished protection from vaccination, despite being capable of maintaining their antibody responses to this virus (9,17). In HD patients, seropositivity ranged from 34.4% to 62.2% six-months after their initial two-dose immunisation schedule, consistently lower than healthy-controls regardless of whether their primary vaccination schedule employed the PFZ or the AZ vaccine.

Encouragingly, following a third vaccination to HD and other CEV patients, antibody levels against all three spike proteins significantly increased and overall seroprevalence against each spike protein exceeded 97%. These data provide reassurance that booster immunisations with currently available vaccines induce relevant humoral immunogenicity against the highly transmissible Omicron VOC (18).

We have previously demonstrated that an extended dosing interval improved the immunogenicity of an initial two-dose Pfizer immunisation in healthy individuals (19). We demonstrate that, in healthy controls, a heterologous vaccination strategy (i.e. AZ prime, followed by PFZ booster), induced antibody responses of greater magnitude against the Wuhan and Delta, but not the Omicron VOC. These data are consistent with clinical trial data demonstrating the increased immunogenicity of a heterologous vaccination strategy against the Wuhan spike glycoprotein (20). Waning of antibodies responses against all three spike proteins following a two-dose vaccination schedule appears greater in recipients of the AstraZeneca vaccine. These observations are of relevance to the design and implementation of future vaccination schedules.

We have previously reported increased morbidity and mortality (21) and impaired SARS-CoV-2 vaccine responses (Shields et al manuscript under submission) in a range of immunologically vulnerable individuals. Intact humoral immune responses are an essential component of robust immunity against SARS-CoV-2 (22,23). In individuals who fail to mount an effective humoral response to vaccination, monoclonal antibodies provide a rational prophylactic or therapeutic approach. We provide evidence of the antibody binding activity of the therapeutic monoclonal antibody Sotrovimab (GSK) against all three spike proteins, including Omicron.

Strong correlations exist between antibody binding and neutralisation (24) and between the presence of neutralising antibodies and protection against severe COVID-19 (25). However, it will be important to correlate our findings of antibody binding with neutralisation assays going forward.

In conclusion, we provide evidence for the need for a third dose of vaccination due to a waning antibody response at 6 months and the broadly cross-reactive humoral immunogenicity of the third vaccine dose against rapidly evolving SARS-CoV-2 VOC in healthy and HD patients.

## Data Availability

All data produced in the present study are available upon reasonable request to the authors

## Acknowledgements

We thank the staff and patients that have kindly volunteered for this study. Thanks also to Abingdon Health for the Wuhan and Delta antigen.

The authors would like to acknowledge the COVID-HD Birmingham Study Group and PITCH consortium that have enabled this work, the staff of the Clinical Immunology Service, managed by Tim Plant, who helped process the healthcare worker and haemodialysis samples and Dr Margaret Goodall for her expertise in antibody production and assay development. The authors would also like to acknowledge the National Institute for Health Research (NIHR)/Wellcome Trust Birmingham Clinical Research Facility and University Hospitals Birmingham Research and Development team, in particular the research nurses that enabled the sample consent and sample collection including Mary Dutton, Lesley Fifer, Sinead White, Natalie Walmsley-Allen, Lucy Atchinson-Jones, Kulli Kuningas, Margaret Carmody, Rani Maria Joseph, Christopher McGhee, Shannon Page and Michelle Bates. Also The Ulster Pandemic Study team.

The COCO/PITCH healthcare worker cohort was funded by the United Kingdom Department of Health and Social Care and United Kingdom Research and Innovation COVID-19 Rapid Response Rolling Call as part of the PITCH Consortium. The HD cohort was funded by the United Kingdom Research and Innovation COVID-19 Rapid Response Rolling Call.

The COVID-HD Birmingham Study Group include Claire Backhouse, Anna Casey, Lynsey Dunbar, Beena Emmanuel, Megan Fahy, Alexandra Godlee, Paul Moss, Peter Nightingale, Liz Ratcliffe, Stephanie Stringer, Matthew Tabinor, Sian Faustini, Adam Cunningham, Alex Richter, Lorraine Harper.

The PITCH study Group include Susanna Dunachie, Paul Klenerman, Lance Turtle, Thusan I. de Silva, Christopher Duncan, Rebecca Payne, Alex Richter, Ellie Barnes, Miles Carroll, Alexandra Deeks, Christina Dold.

## Figure legends

Supplementary Figure 1

A – Standard curve of NIBSC 20-136 standard against Wuhan, Delta and Omicron anti spike IgG. NIBSC – National Institute for Biological Standards and Controls.

B – Dilution series of Sotrovimab monoclonal antibody therapy against Wuhan, Delta and Omicron anti spike IgG.

## References

1. Voysey M, Clemens SAC, Madhi SA, Weckx LY, Folegatti PM, Aley PK, et al. Safety and efficacy of the ChAdOx1 nCoV-19 vaccine (AZD1222) against SARS-CoV-2: an interim analysis of four randomised controlled trials in Brazil, South Africa, and the UK. Lancet. 2021 Jan;397(10269):99–111. doi: 10.1016/S0140-6736(20)32661-1

2. Polack FP, Thomas SJ, Kitchin N, Absalon J, Gurtman A, Lockhart S, et al. Safety and Efficacy of the BNT162b2 mRNA Covid-19 Vaccine. N Engl J Med. 2020 Dec;383(27):2603–15. doi: 10.1056/nejmoa2034577

3. Davies NG, Abbott S, Barnard RC, Jarvis CI, Kucharski AJ, Munday JD, et al. Estimated transmissibility and impact of SARS-CoV-2 lineage B.1.1.7 in England. Science (80-). 2021; doi: 10.1126/science.abg3055originally

4. WHO World Health Organisation. Tracking SARS-CoV-2 variants. ONLINE. 2021.

5. Science Brief: Omicron (B.1.1.529) Variant. CDC COVID-19 Sci Briefs. 2020; [accessed 23 Dec 2021] Available from: https://pubmed.ncbi.nlm.nih.gov/34932278/

6. Shields AM, Faustini SE, Kristunas CA, Cook AM, Backhouse C, Dunbar L, et al. COVID-19: Seroprevalence and Vaccine Responses in UK Dental Care Professionals. J Dent Res. 2021 Jun;220345211020270. doi: 10.1177/00220345211020270

7. Shields A, Faustini SE, Perez-Toledo M, Jossi S, Aldera E, Allen JD, et al. SARS-CoV-2 seroprevalence and asymptomatic viral carriage in healthcare workers: a cross-sectional study. Thorax. 2020 Sep 7;thoraxjnl-2020-215414. Available from: http://thorax.bmj.com/content/early/2020/08/28/thoraxjnl-2020-215414.abstract

8. Shields AM, Faustini SE, Perez-Toledo M, Jossi S, Allen JD, Al-Taei S, et al. Serological responses to SARS-CoV-2 following non-hospitalised infection: Clinical and ethnodemographic features associated with the magnitude of the antibody response. BMJ Open Respir Res. 2021 Sep 24;8(1). doi: 10.1136/bmjresp-2020-000872

9. Banham GD, Godlee A, Faustini SE, Cunningham AF, Richter A, Harper L. Hemodialysis patients make long-lived antibodies against sarscov-2 that may be associated with reduced reinfection. J Am Soc Nephrol. 2021 Sep 1;32(9):2140–2. doi: 10.1681/ASN.2021020188

10. Faustini SE, Jossi SE, Perez-Toledo M, Shields AM, Allen JD, Watanabe Y, et al. Development of a high-sensitivity ELISA detecting IgG, IgA and IgM antibodies to the SARS-CoV-2 spike glycoprotein in serum and saliva. Immunology. 2021 Sep 1;164(1):135–47. doi: 10.1111/imm.13349

11. Cook AM, Faustini SE, Williams LJ, Cunningham AF, Drayson MT, Shields AM, et al. Validation of a combined ELISA to detect IgG, IgA and IgM antibody responses to SARS-CoV-2 in mild or moderate non-hospitalised patients. J Immunol Methods. 2021 Jul 1;494. doi: 10.1016/j.jim.2021.113046

12. Watanabe Y, Allen JD, Wrapp D, McLellan JS, Crispin M. Site-specific glycan analysis of the SARS-CoV-2 spike. Science. 2020 Jul 17;369(6501):330–3. Available from: https://pubmed.ncbi.nlm.nih.gov/32366695/

13. NIBSC NI for BS and Controls. WHO International Standard First WHO International Standard for anti-SARS-CoV-2 immunoglobulin (human) NIBSC code: 20/136 Instructions for use (Version 2.0, Dated 17/12/2020). Potters Bar, Hertfordshire, EN6 3QG;

14. Fabian Schmidt, Frauke Muecksch, Yiska Weisblum, Justin Da Silva, Eva Bednarski, Alice Cho et al. Plasma neutralization properties of the SARS-CoV-2 Omicron variant. MedRxiv 2021 Dec 13;2021.12.12.21267646. Preprint. PMID: 34931199

15. Wilfredo F Garcia-Beltran, Kerri J St Denis, Angelique Hoelzemer, Evan C Lam, Adam D Nitido, Maegan L Sheehan, et al. mRNA-based COVID-19 vaccine boosters induce neutralizing immunity against SARS-CoV-2 Omicron variant. MedRxiv. 2021 Dec 14;2021.12.14.21267755. Preprint. PMID: 34931201

16. SARS-CoV-2 Omicron has extensive but incomplete escape of Pfizer BNT162b2 elicited neutralization and requires ACE2 for infection. Sandile Cele, Laurelle Jackson, Khadija Khan, David S Khoury, Thandeka Moyo-Gwete et al. MedRxiv. 2021 Dec 11;2021.12.08.21267417. Preprint. PMID: 34909788

17. Edward J Carr, Mary Wu, Ruth Harvey, Emma C Wall, Gavin Kelly et al. Haemodialysis COVID-19 consortium; Crick COVID Immunity Pipeline;et al. Neutralising antibodies after COVID-19 vaccination in UK haemodialysis patients. Lancet. 2021 Sep 18;398(10305):1038–1041. doi: 10.1016/S0140-6736(21)01854-7. Epub 2021 Aug 13.

18. Pascarella S, Ciccozzi M, Bianchi M, Benvenuto D, Cauda R, Cassone A. The Electrostatic Potential of the Omicron Variant Spike is Higher than in Delta and Delta-plus Variants: a Hint to Higher Transmissibility? J Med Virol. 2021 Dec 16; Available from: https://pubmed.ncbi.nlm.nih.gov/34914120/

19. Payne RP, Longet S, Austin JA, Skelly DT, Dejnirattisai W, Adele S, et al. Immunogenicity of standard and extended dosing intervals of BNT162b2 mRNA vaccine. Cell. 2021 Nov 11;184(23):5699-5714.e11. Available from: https://pubmed.ncbi.nlm.nih.gov/34735795/

20. Munro APS, Janani L, Cornelius V, Aley PK, Babbage G, Baxter D, et al. Safety and immunogenicity of seven COVID-19 vaccines as a third dose (booster) following two doses of ChAdOx1 nCov-19 or BNT162b2 in the UK (COV-BOOST): a blinded, multicentre, randomised, controlled, phase 2 trial. Lancet (London, England). 2021 Dec;398(10318):2258–76. Available from: https://pubmed.ncbi.nlm.nih.gov/34863358/

21. Shields AM, Burns SO, Savic S, Richter AG, Anantharachagan A, Arumugakani G, et al. COVID-19 in patients with primary and secondary immunodeficiency: The United Kingdom experience. J Allergy Clin Immunol. 2021 Mar 1;147(3):870-875.e1. Available from: https://pubmed.ncbi.nlm.nih.gov/33338534/

22. Brown LAK, Moran E, Goodman A, Baxendale H, Bermingham W, Buckland M, et al. Treatment of chronic or relapsing COVID-19 in immunodeficiency. J Allergy Clin Immunol. 2021; Available from: https://pubmed.ncbi.nlm.nih.gov/34780850/

23. Lucas C, Klein J, Sundaram ME, Liu F, Wong P, Silva J, et al. Delayed production of neutralizing antibodies correlates with fatal COVID-19. Nat Med. 2021 Jul 1;27(7):1178–86. Available from: https://pubmed.ncbi.nlm.nih.gov/33953384/

24. Earle KA, Ambrosino DM, Fiore-Gartland A, Goldblatt D, Gilbert PB, Siber GR, et al. Evidence for antibody as a protective correlate for COVID-19 vaccines. Vaccine. 2021 Jul 22;39(32):4423–8. Available from: https://pubmed.ncbi.nlm.nih.gov/34210573/

25. Khoury DS, Cromer D, Reynaldi A, Schlub TE, Wheatley AK, Juno JA, et al. Neutralizing antibody levels are highly predictive of immune protection from symptomatic SARS-CoV-2 infection. Nat Med. 2021 Jul 1;27(7):1205–11. Available from: https://pubmed.ncbi.nlm.nih.gov/34002089/

