## Supplementary figures and images for "Cross reactivity of spike glycoprotein induced antibody against Delta and Omicron variants before and after third SARS-CoV-2 vaccine dose"

### Supplemental Figure 1a &1b

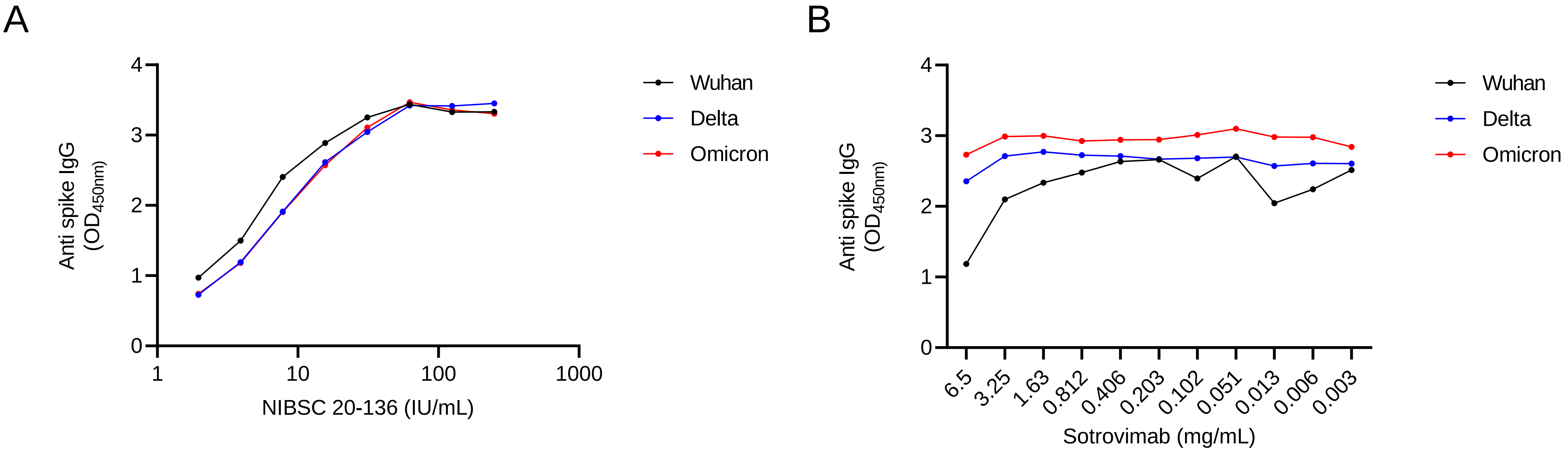
